# EpiControl: a data-driven tool for optimising epidemic interventions and automating scenario planning to support real-time response

**DOI:** 10.1101/2025.11.17.25340271

**Authors:** Sandor Beregi, Sangeeta Bhatia, Anne Cori, Kris V. Parag

**Affiliations:** MRC Centre for Global Infectious Disease Analysis, Imperial College London, London, UK; Department of Engineering, King’s College London, London, UK

**Keywords:** public health decisions, non-pharmaceutical interventions, vaccination, real-time, scenario analysis, outbreak preparedness

## Abstract

Deciding among outbreak control policies is challenging because available data are imperfect and intervention effects are uncertain. Complex disease models can simulate the economic and health consequences of interventions, but substantial data or computational demands coupled with the specialist knowledge needed for calibration limit their real-time application. Simpler, faster and more generalised tools exist but typically they only estimate or forecast transmission dynamics under preset assumptions. We present EpiControl, a flexible tool (in R) for public health modellers to support policymaking that fits semi-mechanistic models to routine data and leverages feedback-control and model-based learning to balance complex policy-simulation with real-time responsiveness. EpiControl generates intervention scenarios that automatically update with unfolding dynamics, minimise costs, meet user-defined policy targets (e.g., suppressing epidemic peaks) and remain robust to unanticipated changes. Using Ebola virus and COVID-19 case studies, we establish how EpiControl can rapidly discover optimal intervention policies that prevent hospital overload, reduce societal disruption and retain control despite uncertain immunity, vaccination and pathogen variant dynamics.

## Introduction

The effective control and management of infectious disease outbreaks is a principal challenge that demands timely, data-driven and adaptive public health intervention policies [1], [2]. Deciding on how and when to initiate, intensify, switch, relax or remove control measures is a problem with multiple and often conflicting objectives that is subject to social, economic and biological constraints together with uncertainties in surveillance, transmission dynamics and intervention outcomes [1], [3], [4], [5]. During unfolding epidemics, the need for evidence-based decision tools is paramount, both for balancing actions with uncertainty and risk, amidst dynamically growing infections and because the cost and benefits of those actions are highly sensitive to implementation and timing [6], [7], [8], [9]. As examples, model-based outputs were critical for allocating resources to control the 2015 Ebola virus epidemic [10], where the high disease fatality rate amplified the risks of delays and were instrumental for informing the initiation and removal of a suite of non-pharmaceutical interventions (NPIs) throughout the 2020 COVID-19 pandemic [11], in the face of enormous uncertainties about the dynamics of a novel, emerging coronavirus.

Historically, a key approach to designing, selecting and testing control policies has relied on parameterising and running complex epidemiological and socio-economic decision models [11], [12], [13], [14]. These realistically simulate the mechanisms that underpin transmission (e.g., contact networks) and provide detailed projections and assessment of how interventions can disrupt those mechanisms to regulate spread. Although invaluable for informing policy, these models can often be difficult to interrogate, interpret, validate and fit without extensive data, intensive compute and specialist knowledge about the disease of interest. Moreover, the effectiveness of actions is sensitive to implementation details and likely to vary as the epidemic evolves [6], [15], [16]. These factors all limit the timeliness and usefulness of this approach for *real-time* responses due to scarce data as well as high uncertainty around pathogen traits or intervention effects and because of a need to repeatedly refit models to data as the outbreak unfolds [17], [18].

Recognising these drawbacks, alternative and simpler modelling approaches have been used to guide real-time preparedness and response. These typically involve parsimonious, semi-mechanistic transmission models that are fit to routinely available data to provide continuously updating estimates of key parameters such as time-varying reproduction numbers and growth rates [19], [20], [21]. This minimal set of interpretable parameters is frequently sufficient to generate projections of epidemic burden, assess intervention outcomes and inform decision-making [22], [23], with EpiEstim as one of the most established of these approaches [19]. While these methods are crucial for situational awareness of the epidemic state, when applied to scenario analysis they take what is termed an *open loop* perspective [24], [25], [26], [27], [28], operating under preset assumptions, for example simulating epidemic outcomes under predetermined intervention schedules, strengths and durations. Typically, these approaches do not allow for feedback loops, where proposed interventions update with the epidemic trajectory, limiting the ability to respond to changing disease characteristics or refine counterfactual scenarios.

Moreover, across both simpler and complex models there has been a growing call for including formal decision-analysis and feedback against clearly defined target objectives within scenario planning [4], [5], [15], [29], [30]. This is the *closed loop* perspective and provides frameworks not only for rigorously comparing interventions and counterfactual outcomes but also for adapting policies to dynamic disease landscapes and unmodelled phenomena (e.g., unanticipated population behaviours). This adaptive property derives from corrective feedback between the epidemic state (inferred from data) and desired targets or objectives (set by policymakers). Although the closed loop perspective is starting to shape epidemiological thinking [24], [29], [31], [32], [33], a flexible scenario analysis tool bridging the optimisation and counterfactual approaches of complex models with the real-time effectiveness of minimalistic methods such as EpiEstim is lacking.

Here we develop and validate an automated, data-adaptive, scenario planning tool in R called EpiControl to fill this gap. EpiControl intakes user-defined targets (e.g., on the peak or endemic infection levels), routine data (e.g., incidence time series of epidemiological events) as well as estimates of the relative costs and efficacies of interventions (defined broadly) and leverages algorithms from feedback control and model-based reinforcement learning to rapidly identify closed-loop control policies that meet those targets. These policies maximise long-term value and improve robustness to uncertainties and unmodelled dynamics, even when input costs and efficacies are poorly specified. This robustness and adaptability stem from refocussing scenario design on explicit target objectives and using errors relative to targets to dynamically adjust policies in real time. EpiControl helps find how, when and if public health objectives can be realised.

We start by outlining the methodology underpinning EpiControl and its usage with real data. We then demonstrate performance on a range of empirical and simulated data including those underlying influential studies that provided valuable NPI (non-pharmaceutical intervention) and vaccine policy recommendations across the COVID-19 pandemic [11], [34]. We find that EpiControl provides fast, optimal and dynamic control policy suggestions that adapt to emerging changes (e.g., pathogenic variants) and uncover solutions within the decision space that may be overlooked by standard approaches. We benchmark the performance of EpiControl (via sensitivity tests and measures of computational complexity) in the Supplement.

## Methods Overview

### The EpiControl scenario and counterfactual analysis tool

EpiControl is an open-source, scenario simulation R-package that frames outbreak decision-making as a real-time, adaptive control problem and enables epidemic modellers that inform policymakers to rapidly explore counterfactual scenarios and assess the likely consequences of interventions ahead of time. EpiControl applies receding-horizon, model predictive control (MPC) to discover optimal interventions [35]. This is equivalent to a model-based reinforcement learning approach [36], except that policies are not trained offline [37]. Instead, they are planned, executed and re-optimised sequentially in real-time. With EpiControl, users can rapidly explore and exploit optimal policies to support timely decisions or guide more complex modelling.

We outline the modular structure of EpiControl in **Fig 1**. There are 5 plug-and-play modules: a model of the ongoing disease outbreak, a real-time estimator of transmissibility, a projection model with a library of candidate actions (e.g., various NPI choices), a user-defined cost function with elements considering the disruption caused by NPIs and penalties for violating user targets or increasing epidemic burden and a model of surveillance imperfections that distorts the true epidemic data into practical observations. The value function of a chosen intervention is the cumulative cost over stochastic projections of the epidemic under that intervention and factors in the longer-term rebound or feedback effects of NPI choices. Costs and value functions are re-evaluated at every policy review time (e.g., every two weeks or monthly). EpiControl generates NPI scenarios that meet policy targets at minimal cost or reveals when our library of NPIs cannot achieve those targets (evidencing when new interventions including pharmaceuticals are needed).

**Fig 1:**
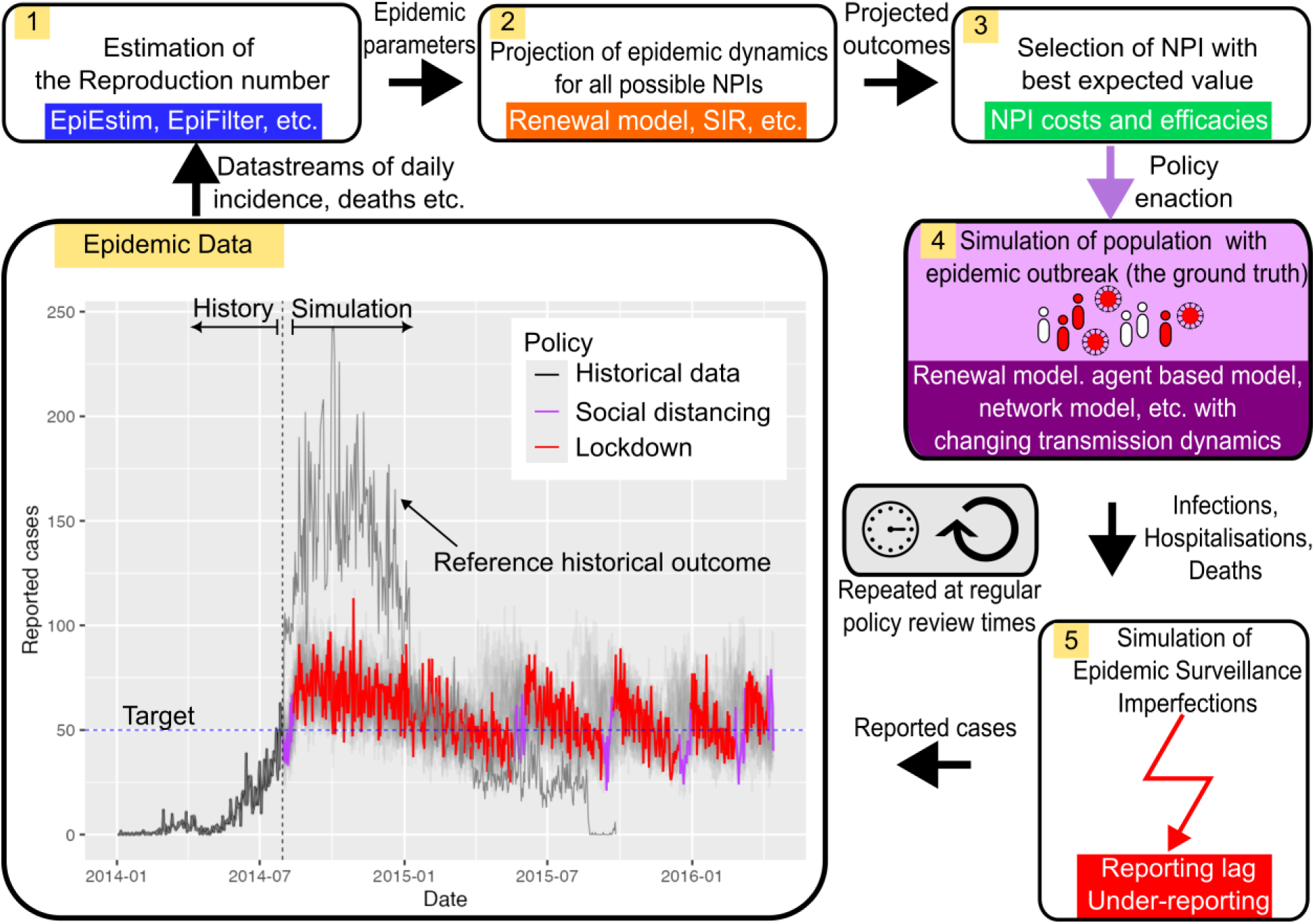
The EpiControl simulation and policy optimisation framework with an Ebola virus example. The Epidemic Data panel shows reported case incidence from the 2014–15 West African Ebola virus outbreak, with historical empirical data shown in black. The vertical dashed line separates the observed historical period from the simulated control period. The horizontal dotted line is the target daily Ebola virus case counts, around which the user wants to stabilise the epidemic in this example. Grey trajectories show stochastic reference outcomes, while the highlighted controlled trajectory shows the selected adaptive policy. Purple and red sections denote intervention tiers, with red representing the more stringent lockdown policy. We summarise the iterative EpiControl workflow using numbered panels. In panel 1, surveillance streams, such as reported cases, deaths or hospitalisations, are used to estimate epidemic parameters, including the time-varying reproduction number. In panel 2, these estimates are passed to a forecasting model, such as a renewal, compartmental, agent-based or network model, to project epidemic dynamics under all candidate non-pharmaceutical interventions (NPIs). In panel 3, EpiControl evaluates projected outcomes using user-defined intervention costs, efficacies, and epidemic targets, and selects the NPI with the highest expected value. In panel 4, the selected policy is enacted in the simulated population, generating infections, hospitalisations or deaths under the underlying epidemic dynamics. In panel 5, imperfect surveillance observations are produced, including simulated reporting delays and under-reporting. These observations are fed back into panel 1 at the next scheduled policy review time, producing an adaptive policy revision cycle that can respond to changing transmission dynamics and surveillance uncertainty. Each panel represents a modular, customisable component of EpiControl. Within each panel, in the coloured boxes, we provide examples of custom methods or functionalities that can be used within that module.

We model the incidence of new infections as a stochastic renewal process. This models new infections as Poisson distributed with a mean that depends on past infections, the time-varying reproduction number *R_t_*, which is the average number of new infections generated by an infectious individual at *t* and the disease generation times, which are the random durations between infections [19]. We also include real-world surveillance noise sources such as reporting delays and under-ascertainment, which distort the true infection incidence into commonly observed proxies such as the incidence of cases. We provide details of all these transmission and noise models in the Methods. Our default model is parsimonious to prioritise the rapid evaluation of numerous scenarios, but any transmission model outputting daily case incidence can, in principle, be substituted and the same EpiControl algorithms applied.

Users set a realistic schedule for policy revisions, e.g., every 2-4 weeks. This feature reflects the practicalities of public health interventions: policies are generally only updated infrequently and there are delays between announcing and realising actions. At the end of each review period, EpiControl revises its estimate of the time-varying reproduction number *R*_t_ from the additional data that emerged since the last review and uses it to infer a baseline reproduction number. EpiControl then searches across a library of candidate NPIs (supplied by the user e.g., from studies like [38]) with each NPI parametrised by a relative cost and a multiplicative reduction to the baseline reproduction number. These cost and effect parameters can be fixed or modelled probabilistically as set by users and can be updated across time. This allows inclusion of uncertainties and flexible testing of counterfactual NPI scenarios.

At every policy review, EpiControl projects epidemic trajectories under candidate NPIs over a time-horizon. This horizon balances the need to capture the intervention effect, which may take more than a generation to be fully observed, against growing uncertainty from forecasts. Over every projection, a value function that embeds policy priorities or targets such as limiting adverse health outcomes (e.g., from big epidemic peaks) and economic disruption from NPIs (e.g., costs that scale with stringency and duration of the control action) is evaluated. Across our analyses, we use lockdown to represent to the most stringent NPI (highest transmission reduction and implementation cost). We term intermediate tiers of interventions as social distancing. We apply a discount factor when compounding value over the horizon to reflect increasing uncertainties from longer-term predictions. The NPI with the largest discounted value, averaged across an ensemble of realisations, is implemented, closing the feedback loop between data and policy action. Repeating this process across review times provides adaptability and robustness.

The value function and discount factor are fully customisable to align EpiControl scenarios with local priorities, heterogeneities and risk profiles. The algorithms underpinning EpiControl derive from receding horizon or model predictive control and reinforcement learning theory and extend approaches from [25] to improve robustness, uncertainty handling, account for changes in susceptibility (from infections or vaccinations) and incorporate multiple streams of data. We tune any free parameters (e.g., horizon lengths) using Bayesian optimisation and provide algorithms to tailor tuning to diseases of interest. We provide examples of this tuning as well as of 4 key case studies as online vignettes in the EpiControl package. These case studies are explored in the Results and illustrate the broad functionality of EpiControl.

Case study 1 establishes how EpiControl can outperform the threshold-based schemes from Report 9 by the Imperial College London COVID-19 Response Team [11], which were highly impactful in guiding global COVID-19 policy. We find new schemes that improve peak and total intensive care unit (ICU) cases with the same level of intervention usage. Case study 2 considers Delta variant emergence during the vaccination rollout against COVID-19 in the UK. The Roadmap out of Lockdown [39], [40] was developed to test scenarios in response, but at the time, no tool was available that could automatically identify optimal intervention policies and adapt them to unforeseen, unmodelled changes in transmission, such as the emergence of the more transmissible Delta variant or uncertainty in vaccination uptake. EpiControl demonstrates how such adaptive planning could generate policies that remain effective under these changing conditions. In contrast, the existing policy was unable to manage the Delta variant reliably. Case studies 3-4 showcase the generalisability and reliability of EpiControl, examining how noise from different data streams limits how well we can minimise Ebola virus disease and COVID-19 deaths and exploring how uncertainty over NPI libraries and effect sizes shape optimal exit strategies as we approach the herd immunity threshold.

## Results

### 1. Dynamically optimising NPI responses to emerging epidemics (Report 9)

Report 9 [11] was a highly impactful study in early 2020 that modelled NPI scenarios for the then emerging COVID-19 pandemic. It examined whether combinations of NPIs could prevent ICU cases, caused by COVID-19 infections, from exceeding healthcare capacities in the UK. Report 9 applied an epidemiologically detailed, computationally complex, agent-based model and proposed recurrent NPI strategies that were triggered when weekly ICU cases exceeded a threshold of 100 and lifted when they subsided below 50. This (implicit) feedback-control policy cycled restrictions using observed epidemic data and outperformed key counterfactuals at that time: a full suppression strategy that would be excessively disruptive and a milder policy restricting the epidemic to one peak (to prevent later waves), which would overload healthcare capacities. Report 9 informed UK policy and was cited in official documents from government [41], [42].

However, this thresholding strategy has some important drawbacks. By using ICU case counts as thresholds, actions are constrained by the lag between COVID-19 infection and admission to ICU. This meant that, relative to dynamics of transmission, control was delayed, leading to large peaks of ∼400 weekly ICU cases (a 300% overshoot of the on trigger). Further, the policy was inflexible as thresholds were fixed and the design process did not optimise explicit policy targets or objectives. Here we demonstrate how the deployment, efficiency, transparency and potential benefit of this strategy can be substantially improved without increasing NPI usage by using adaptive approaches such as EpiControl. We establish that EpiControl allows rapid optimisation of when and how to introduce the NPIs and makes interpretable what targets are driving these variations. We discover solutions that reduce total and peak ICU cases or reduce the number of NPI actions, opening trade-offs that may help guide the use of more complex models as in [11].

We start by calibrating a homogeneous renewal model to approximate the salient dynamics of the agent-based model from Report 9. We used COVID-19 parameters from [11] including an infection to ICU admission rate of 1.32%, a basic reproduction number R_0_ of 2.2 and mean generation time of 6.5 days. In line with [11], we assume that strong restrictions, which we term “lockdown”, reduce the reproduction number to 0.66. **Fig2a** illustrates that our renewal model is well calibrated as we recover very similar dynamics to that from Report 9 (apart from the first peak which we do not count) when we apply almost the same policy. Our thresholds are 98 and 49 closely reproducing 100 and 50 in Report 9. These are illustrative examples and not actual recommendations.) We run EpiControl with this calibrated model to optimise the implementation and removal of lockdown but according to explicit objectives such as reducing ICU peaks and keeping baseline ICU admissions near a target steady state between the thresholds.

We find that, using the same ICU control signal as [11] and a realistic policy review time of 2 weeks i.e., EpiControl recomputes strategies every 14 days, we can shrink peaks by over 25% to 200-300 weekly ICU cases as in **Fig 2b**. Moreover, we reduce total ICU cases across the simulated period of 530 days (averaged across 100 stochastic realisations of the epidemic) by 20% (∼1700 fewer cases) as compared to **Fig 2a**. We plot this in **Fig 2d**. These advantages emerge from the model-predictive optimisation in **Fig 1**. Unlike the Report 9 strategy, which acts only on short-term information, EpiControl plans over a horizon (see Methods), factoring in longer-term information and partially circumventing the delays from basing actions on ICU cases. Our optimisation against explicit targets ensures control decisions prioritise the aspects of the epidemic trajectory that users (or policymakers) deem most valuable.

**Fig 2:**
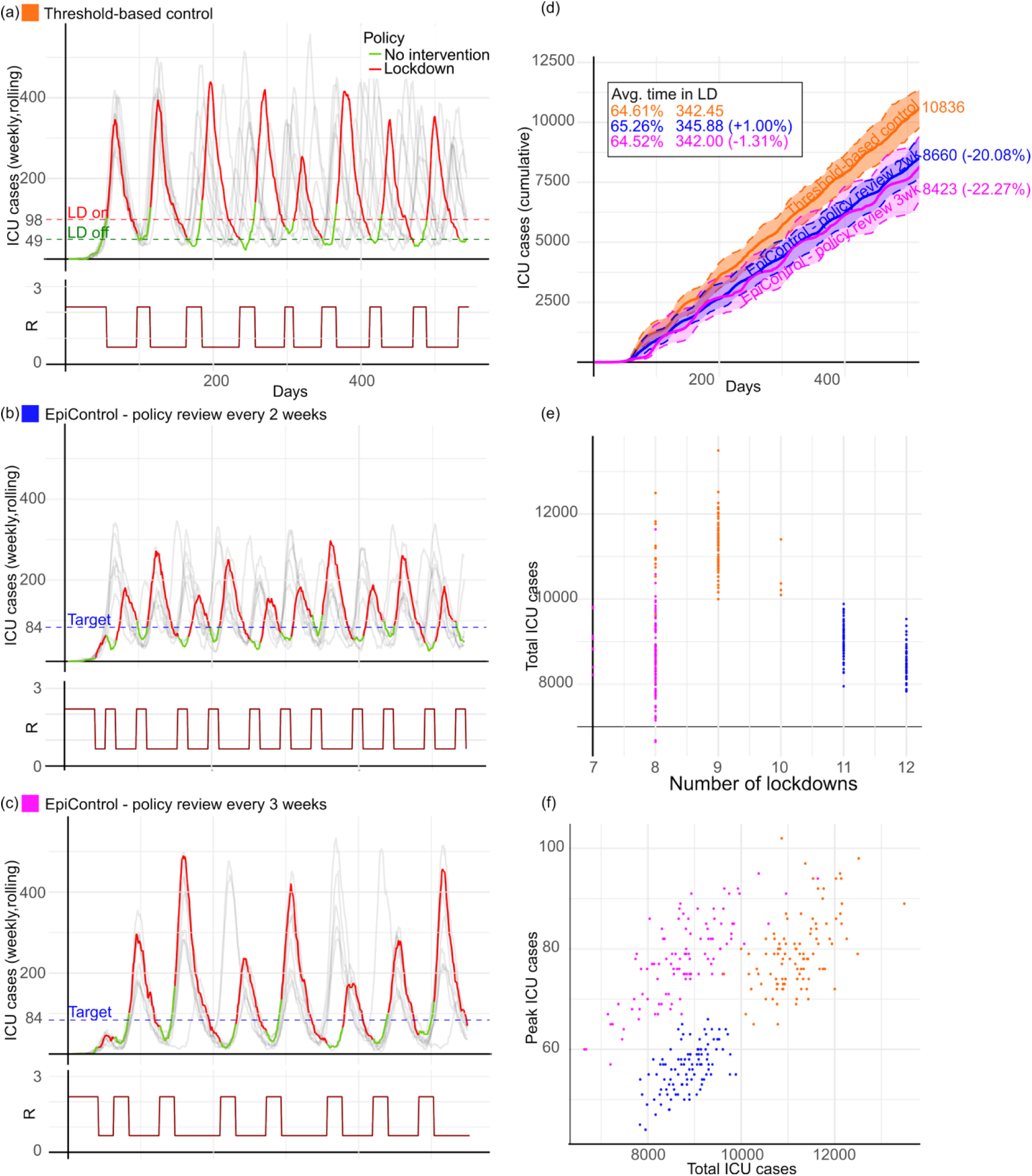
Controlling COVID-19 intensive care unit (ICU) admissions in the UK. We compare optimal EpiControl strategies with a threshold-based policy emulating that proposed in Report 9 [11]. In panel (a), we illustrate the approach of [11], which initiates and ends lockdowns using fixed thresholds (left y axis plots of ICU case trends). Grey thin curves represent different controlled epidemic realisations. We highlight and colour one realisation, with green for no intervention and red showing time spent in lockdown. Red and green dashed lines demarcate on and off thresholds or baseline targets. Below the main panel we plot reproduction numbers R underlying the interventions in the highlighted scenario. Higher R occurs for periods of no interventions and lower R for lockdowns (values from [11]). In panel (b), we apply EpiControl with a target baseline set between the on/off triggers of (a). This shrinks ICU peaks by at least 25% at the expense of 2-3 more lockdowns, but the total time spent in lockdown is unchanged. We consider an alternative control design in panel (c) that delays policy reviews in EpiControl from every 2 (as in b) to 3 weeks. The lockdown number now falls below that of the threshold policy in (a). Panels (d-f) summarise intervention outcomes across realisations. Panel (d) indicates the median and 25-75 percentile range for every method and shows that EpiControl reduces mean total ICU admissions by 20% for the strategy in (b) and 22.3% for the strategy in (c), relative to the threshold policy. Panels (e) and (f) demonstrate trade-offs between total ICU admissions, lockdown frequency and peak ICU admissions, mapping a design space that can inform policymaking. Dots indicate individual realisations and clusters identify strategies that improve on the Report 9-style policy without increasing total lockdown duration.

Importantly, these improvements do not increase the overall duration of time spent under lockdown. However, **Fig 2b** does apply more frequent lockdowns (11–12 versus the 8–10 from **Fig 2a**). It is generally hypothesised in studies on COVID-19 NPIs that both the length and frequency of restrictions (or breaks between stricter NPIs) can change NPI efficacies due to behavioural factors such as adaptation, compliance or fatigue [43]. Some policymakers may therefore prefer different trade-offs among ICU cases averted and the number of lockdowns initiated. EpiControl allows rapid examination of the intervention space to discover effective compromises and new solutions. In **Fig 2c** we extend the policy review time to 3 weeks. As a result, EpiControl reduces the lockdown number (7–8) to fewer than that of the Report 9 strategy. As the total time spent in lockdown is conserved, each lockdown is of longer duration. Interestingly, this policy opens a new trade-off. It achieves a similar peak to **Fig 2a** but markedly reduces the total ICU cases by ∼22%. We note that the policies do not differ at the very start of the simulations because all scenarios are initialised with 10 infections and 0 ICU cases. Before ICU occupancy approaches either the control target or the lockdown-on threshold, none of the controllers has any reason to introduce lockdown. This is effectively a pre-intervention phase in which no algorithm acts. We therefore focus on total simulated outcomes when assessing policy performance.

**Fig 2** and **Fig 2f** present the trade-offs achievable by all 3 strategies (the Report 9 approach and the 2- and 3-week EpiControl policies) providing a decision-space that policymakers or modellers informing policy can use to quantitatively assess strategies of interest. Because we can easily modify, re-parametrise and refit EpiControl against tailored policy priorities, it offers a systematic means of testing counterfactual scenarios ahead-of-time. For example, panels (e)-(f) reveal (for the first time to our knowledge) that it is possible to optimise total ICU cases and either the number of lockdowns or peak ICU cases relative to the policy of [11] by better timing and distributing the same NPIs. This is valuable as studies [7], [25] have found that delays in routine surveillance data can make it difficult to identify the need for NPIs early enough for timely implementation.

The flexible, real-time scenarios that EpiControl provides can augment model-based decisions or guide how complex models such as that in Report 9 are used, accelerating optimal policy discovery. To provide confidence in these optimal solutions, in the Supplementary Material (**Fig S1**) we test the sensitivity of the epidemic outcomes (peak and cumulative ICU cases) to small variations in the control target. We find that controlled results are robust to small target variations and do not lead to abrupt or disproportionate changes in public-health outcomes.

### 2. Robust scenario planning (unexpected variants and a vaccine-based Roadmap)

Case study 1 assumed (as in Report 9) known disease dynamics and provided scenarios that did not consider unmodelled and exogeneous shifts to transmission and other parameters. In reality, environmental, ecological, behavioural and other dynamics that are difficult to monitor and model subject epidemics to numerous uncertainties and unanticipated changes. The UK COVID-19 vaccination programme in 2021 presents an important real-world example. After an initial lockdown in early 2021, the UK government produced a Roadmap out of Lockdown [40] for England that modelled vaccine rollout schedules and considered how expected rises in immunity allow for a tiered, gradual relaxation of NPIs, with less stringent measures enacted until restrictions are no longer required. However, these planned relaxations had to be redone later in 2021 because the more transmissible and vaccine evasive Delta variant emerged and eventually supplanted the originally circulating Alpha variant [34], [39].

Here we demonstrate how EpiControl, due to its combined feedback and learning algorithms, can automatically adapt its proposed intervention schedules to unexpected changes, making it easier to provide robust scenario plans. Using empirical case data and model parameters from [44], we first show in **Fig 3** that EpiControl supports the Roadmap assuming there is no Delta variant emergence (left panels). We set a target of stabilising daily cases around 5,000 and run EpiControl from the first vertical dashed timepoint. **Fig 3a** applied a lockdown until we are below this target then lifts it before successively introducing weaker NPIs. **Fig 3c** plots the vaccination and immunity build-up (which EpiControl is unaware of), while **Fig 3e** shows how the NPI choices change the reproduction number *R_t_* over time. Alpha has a basic reproduction number *R_0_* of 2.61 and the vaccine an efficacy of 83% [44].

**Fig 3:**
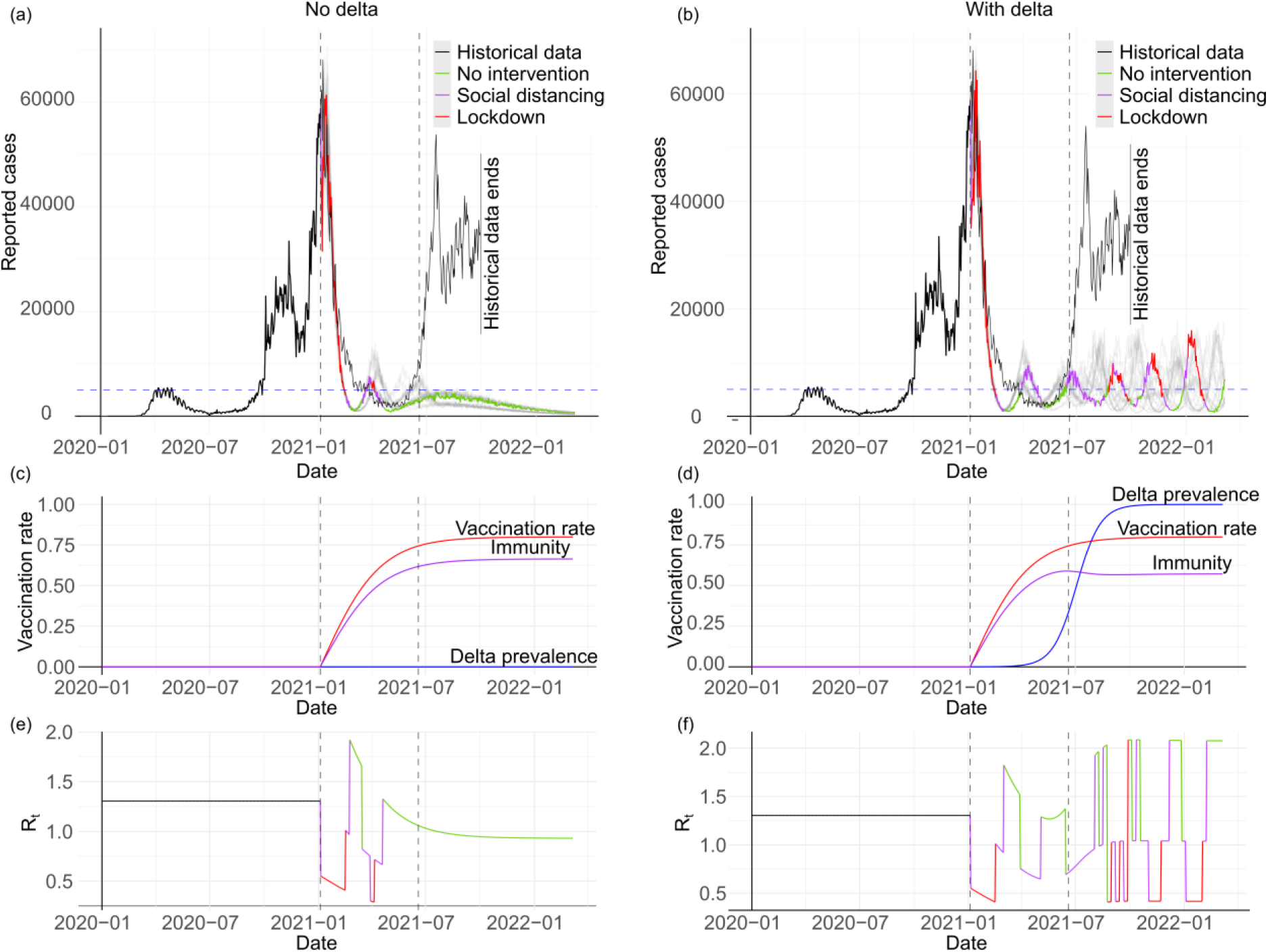
Robust COVID-19 interventions despite unknown vaccination efficacies and Delta variant emergence. Panels (a)-(b) show daily symptomatic cases (solid black, sourced from [45] for the time period between 1 January 2020 and 1 October 2021). We initialise the simulated trajectories with historical data until 1 January 2021 (dashed vertical line) after which EpiControl proposes intervention scenarios coloured by the control measures suggested. In (a) the currently circulating Alpha variant is assumed, while in (b) the more contagious Delta variant emerges and eventually dominates. Delta has 75% higher transmissibility than Alpha and vaccine efficacies are 83% and 71.5% for Alpha and Delta respectively [34]. Our control algorithm is unaware of these values and has no knowledge of panels (c)-(d), which track the proportions of the population that are vaccinated and immune as well as the relative prevalence of Delta (modelled with a logistic function). The vertical dashed line around July highlights when the herd immunity threshold for Alpha is achieved. In the Delta scenario panels, this is provided for reference as overall herd immunity is not achieved in the timeframe examined due to the reduced vaccine efficacy against Delta. Panels (e)-(f) depict the expected controlled reproduction number from the proposed control policies. In the Alpha-only scenario of (a), EpiControl enacts and releases a lockdown, approximating what initially happened (in black). Subsequently, EpiControl reduces NPI stringency and removes all NPIs around when herd-immunity is achieved. This aligns with what the UK Roadmap [40] targets as exit strategy out of lockdown and emerges purely from optimising NPIs against the target baseline (horizontal dashed line), while adapting to inferred changes in transmissibility. In the multi-variant scenario, elevated Delta transmissibility and reduced vaccine effectiveness mean that similar control policies will fail and the high level of infections in the real data after July reflect the impact of Delta. However, despite being agnostic to these exogenous factors, EpiControl is still able to generate intervention options that result in a controlled epidemic, reintroducing lockdowns as needed. EpiControl combines the feedback error between the current epidemic trajectory and user-defined targets with updated learning of transmissibility to achieve this adaptation. The marked gap between coloured scenarios and what actually happened (black) suggest that improvements may have been possible under these policy recommendations.

The proposed scenario (coloured by NPI choice, other stochastic realisations in grey) matches the real data initially (the lockdown corresponds to the actual UK lockdown in early 2021) and removes all restrictions after July 2021, which coincides with the herd immunity threshold (second vertical dashed line). A tiered relaxation of NPIs occurs between these points that aligns with the Roadmap. Interestingly, at no point does EpiControl know the vaccine numbers or when herd immunity occurs. Instead, by sequentially optimising (at every 2-week review) against our targets and projecting the impact of actions (with updating reproduction number estimates), EpiControl naturally learns policies that leverage the benefits of the vaccination rollout to minimise overall costs. The target-centric and adaptive nature of EpiControl also provides inbuilt robustness to exogenous factors that impact those targets. We demonstrate this in right panels of **Fig 3**, which consider the Delta variant.

Delta is significantly harder to control because it has an *R_0_* that is 1.75 times that of Alpha and vaccines have a reduced efficacy of 71.5%. Its unexpected emergence and dominance (we plot its relative prevalence and impact on immunity in **Fig 3d**) meant the Roadmap had to be re-evaluated [39] and led to large, sustained case numbers as shown in black. EpiControl, despite not knowing about Delta, recommends scenarios that achieve our target objectives as in **Fig 3b** (coloured, with grey as other stochastic realisations). After the initial lockdown release the growing Delta prevents relaxations and EpiControl proposes additional shorter lockdowns (we show the *R_t_*changes in **Fig 3f**). This meets targets, keeps the epidemic controlled and markedly improves on what happened in reality (black). Within the simulation timeframe herd immunity is not achieved (the dashed line is from Alpha for reference).

Our feedback-learning approach therefore automatically adapts to unanticipated dynamics, reactively refining policies to provide more reliable and transparent (as our target priorities are explicit) policy decision support. Although Delta delayed the relaxation plans of the Roadmap, authorities chose not to reimpose strict NPIs. This spurred the second wave (black) in **Fig 3b**. We can enforce this constraint in EpiControl if desired. However, the fact that EpiControl chose additional lockdowns indicates that it is impossible to achieve our policy targets without strict controls. This highlights a key benefit of our framework: it identifies when suggested policies or constraints may fail and exposes the limits of what is achievable or not ahead of time.

We illustrate the robustness of these results in an extended analysis in the Supplementary Material (see **Fig S2**). There we test performance sensitivity to changing user-targets both for examples with and without Delta variant emergence (as in **Fig 3**), We find near-linear sensitivity, reflecting that our algorithm is able to reliably control the epidemic despite unknown immunity levels and without any dramatic changes to epidemiological trajectories. These results also indicate the flexibility of our approach for exploring policy design spaces.

### 3. The value of different data for informing the control of deaths resulting from infection

Deciding on which data source is most informative for guiding policy decisions is complex and a subject of ongoing debate [46] especially in high-fatality, under-ascertained outbreaks (e.g. those of Ebola virus disease) [47]. Interventions frequently change the transmission properties of the disease, which first manifest in the incidence of new infections. However, we can rarely observe infections and often rely on a proxy such as the incidence of reported cases, deaths or hospitalisations. All are typically under-reported and delayed outcomes of infections. Even if our target objective is to reduce the incidence of that specific outcome, it is unclear which related data source is best for triggering our control decisions. Case study 1 is an instructive example of this issue. Even though EpiControl reduced the peak ICU cases by 25%, large oscillations were unavoidable because lags between infection and ICU admission restrict timely action [25]. Here we demonstrate how EpiControl can support rigorous decisions on what data to analyse for supporting policy.

We consider a common public health priority: how best can we regulate the deaths resulting from infection. We examine this problem in the context of COVID-19 and Ebola virus disease, which have notably different characteristics. COVID-19 has a short mean generation time (6.5 days [48]), is highly transmissible (*R_0_* of 3.5 [49]) and has a relatively small infection-fatality ratio (IFR) of 1% i.e., 1% of infections culminate in mortality [50]. In contrast, Ebola virus transmits more slowly (mean generation time ∼15 days and *R_0_* of 2.5 [25], [51]) but has a large IFR of 50% [52]. We set a (customisable) policy target of keeping mortality near 50 deaths/day for COVID-19 and 100 for Ebola virus and assume that decisions are reviewed, at most, weekly to reflect plausible constraints on real-time policymaking. We test 4 data source scenarios, evaluating the optimal lockdown and social distancing control policies for each scenario with EpiControl in **Fig 4**.

**Fig 4:**
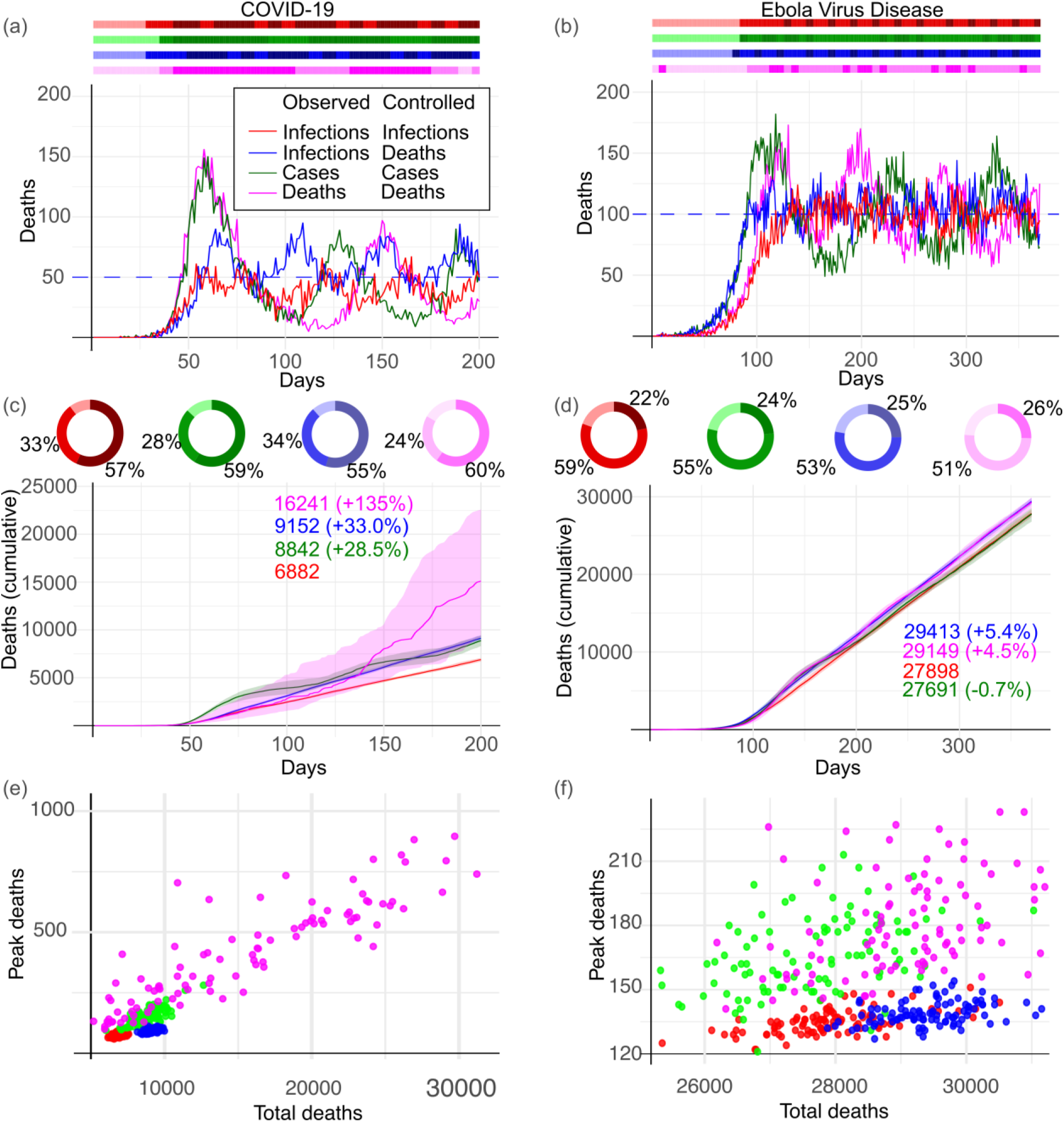
Deciding among data sources to inform the control of COVID-19 and Ebola virus disease death counts. COVID-19 and Ebola virus disease cause large numbers of deaths in different ways. The first has high transmissibility and a relatively low rate of infections leading to death while the second has lower transmissibility and a high infection fatality rate (IFR) (see main text for settings and parameters). We explore how choice of data source for informing NPI decisions impacts our ability to regulate the deaths resulting from each disease around some manageable target. We apply EpiControl to generate optimal policies that use (i) infections assuming we know the IFR (red curves), (ii) infection and death data (blue curves), (iii) reported cases assuming we know the case fatality rate (green curves) and reporting noise (delays and under-ascertainment) and (iv) only death data (magenta curves) i.e., no infection or case data are used. Panels (a) and (b) show the resulting controlled death curves against the target (horizontal, dashed). Bars in the colours of (i)-(iv) indicate how NPIs are deployed with the darkest, medium and lightest tones indicating lockdown, social distancing and no intervention. Panels (c-f) plot performance statistics for scenarios (i)-(iv) in consistent colours collated across 100 epidemic realisations (a and b show one realisation for each strategy). For COVID-19, directly controlling infections as in (i) minimises the cumulative deaths over the study period. Strategies using delayed signals (reported cases or deaths) result in higher peaks and larger mortality. Notably (iii), which infers upcoming deaths from both past deaths and cases, outperforms (iv), which uses only past deaths, drawing benefits from the timelier case reports. In contrast, performance differences among (i)-(iv) are minimal for Ebola virus, as its longer generation time reduces the impact of delays. Pie charts on (c) and (d) display the proportion of time spent under each NPI, which is similar across all scenarios. Consequently, it is the distribution and timing of NPIs (see bar charts in (a) and (b) that distinguish among scenarios (i)-(iv). We plot the design space of intervention outcomes in panels (e) and (f). These confirm that COVID-19 control performance clusters by the type of surveillance data used. For Ebola virus, clustering is less clear, verifying the interaction of surveillance and transmission timescales. EpiControl finds that rapid access to information is crucial for COVID-19 response, but provides comparatively little advantage for Ebola virus, as its slower dynamics make control less sensitive to surveillance delays.

In scenario (i) we observe infections without error and use knowledge of the disease IFRs to design equivalent targets of 5000 and 200 infections/day for COVID-19 and Ebola virus. We therefore control infections around infection targets that should achieve our death targets. This uses the timeliest data for actions and projections. Scenario (ii) uses deaths (reported without error) directly as control signals but applies infection data when performing model projections, isolating the impact of timely signals to assigning the future value of control actions (we do not know the IFRs here). We model the lags between infection and death as Gamma distributions with means of 14.5 [53] and 10 days [54] and standard deviations of 13.82 and 9.53 days respectively for COVID-19 and Ebola virus. Scenario (iii) replaces infections in (i) with reported cases (and uses case fatality ratios), reflecting practical constraints of realistic surveillance, while (iv) assumes we cannot access infections in (ii) and so directly uses past death data to inform decisions and future projections.

Cases are a delayed fraction of infections. We model this delay as a Gamma distribution with mean 10.5 days [55] for both diseases and the reporting fraction as a Beta distribution with a mean of 30% [25]. We recalibrate the target in (iii) to be based on cases using the case fatality ratio (CFR) of both diseases (1% for COVID-19 [56] and 50% for Ebola virus [57]). The death-to-death model fit for scenario (iv) is also a renewal model (mirroring approaches from [58]) with the equivalent of a death-to-death generation. Each of our 4 scenarios present different ways of processing data for controlling deaths and we examine their performance on daily death counts, cumulative mortality (over 100 realisations) and in terms of the distribution (bar charts) and ratio of time spent (pie charts) under our optimal NPI choices in **Fig 4**.

We find that the value of timely information, even when we are interested in a delayed signal (i.e., our death targets) and apply optimal feedback and learning algorithms, depends strongly on implementation and disease characteristics. The timeliness of (i – infection data) outweighs any mismatch arising from stochastic differences between from using implicit infection targets instead of the actual death targets. **Fig 4a**, **4c and 4e** show that COVID-19 control is highly sensitive to data source choice as (ii – infection and death data) already raises total mortality by ∼28% and peak death counts by 200%. Scenario (iii – cases data) helps with the overshoot (as cases are less delayed than deaths) but case reporting noise means ∼33% more deaths than (i – infection data). Scenario (iv – death data only), may seem a reasonable approach (and has been employed during COVID-19) to regulating deaths but is by far the worst signal for control. EpiControl provides clear evidence that even though deaths are more accurately recorded than cases, the earlier detection of transmission trends possible from case data is more important for effective COVID-19 control. The bar and pie charts above **Fig 4a** and **Fig 4b** highlight this as the optimal policies using (i)-(iv) mainly differ in their distribution of NPIs. The total time spent in the various NPIs for all policies is similar.

The value of timely information dramatically drops for Ebola virus disease. **Fig 4b, 4d** and**Fig 4f** illustrate comparable peak and total deaths across the scenarios. Although (iii – cases data) and (iv – death data) have ∼5% more cumulative deaths and higher amplitude oscillations, the importance of choosing among cases, deaths or infections (if available) shrinks. The slower spread of Ebola virus gives policymakers more latitude for choosing control signals and suggests investing in surveillance, for example to reduce reporting delays, may not bring notable improvements in the utility of model-based evidence. This is not obvious given the large IFR of the disease. Accordingly, as ICU cases are between cases and deaths in terms of lag, there is evidence here that surveying more timely signals might improve the optimal polices we found in case study 1.

Interestingly, when we compare individual outcomes over the design space of peak and total deaths in **Fig 4e** and **Fig 4f**, the different strategies show similar tendencies and organise into clusters. Against the baseline strategy of using real-time infection incidence together with the IFR, using deaths as the control signal, even with real-time surveillance, increases total deaths more significantly than peak deaths. In contrast, using reported cases rather than infections primarily increases peak deaths, with a smaller effect on total deaths. Enacting interventions based on deaths without observing the underlying infection incidence is generally the worst-performing strategy, increasing both peak and total deaths. Clusters are less pronounced and more overlapping for Ebola virus because of the closer interplay between its transmission and surveillance timescales.

EpiControl is well suited for informing decisions about which surveillance data to prioritise in relation to policy targets, because it can adaptively model the noise, delays, targets and disease dynamics shaping the reliability of various information sources. This enables it to reveal where further investment in data offers genuine value for improving model-based policy planning and where simpler indicators are sufficient for robust control.

### 4. Complex control with susceptible depletion and multiple, uncertain interventions

The above case studies optimised over limited numbers of interventions, but the landscape of infectious disease control can be very complex with many intervention options that have highly uncertain effects [23]. Optimising over larger intervention spaces can inform decisions among competing and combinatorial actions such as mask mandates, closures, tiered lockdowns, pharmaceuticals and others. Factoring in the uncertainties in intervention effects can enhance the reliability of any trade-offs among those options. In case study 2, for example, we derived optimal strategies for relaxing NPIs as (unknown) population immunity accumulated that were robust to unexpected variant emergence. However, these policies featured oscillations that likely could be improved if intermediate interventions were available. Here we demonstrate how EpiControl can solve arbitrary numbers of interventions with uncertainty in real-time once we specify (to within a distribution) the likely costs and effects of each action.

Using COVID-19 parameters from earlier case studies, we start with a baseline control space in **Fig 5a** that comprises a strict lockdown (effect *c=0.2,* see Methods), an intermediate action we term social distancing (*c = 0.5*) and no restrictions (*c=1*). We apply a weekly policy review time and explicitly model susceptible depletion due to lifelong immunity acquired from infection in a closed population of 3.5 million individuals. Under these conditions the optimal policy first alternates lockdown and social distancing, then switches solely to social distancing before lifting all restrictions once herd immunity has been achieved. EpiControl therefore provides the optimal path to exiting interventions. In **Fig 5d** and **Fig 5f** we expand the action space to include 6 and 10 NPIs, respectively (described in **Fig 5b**). Although this optimisation is harder, EpiControl solves this problem in real time, discovering fine-tuned policies that exploit both the rising immunity and the availability of intermediate and less costly actions to minimise the overall cost until herd immunity. The larger action space does not result in more frequent or drastic policy changes. Instead, weekly control actions are refined gradually and help dampen overshoot and oscillations around the target threshold.

**Fig 5.**
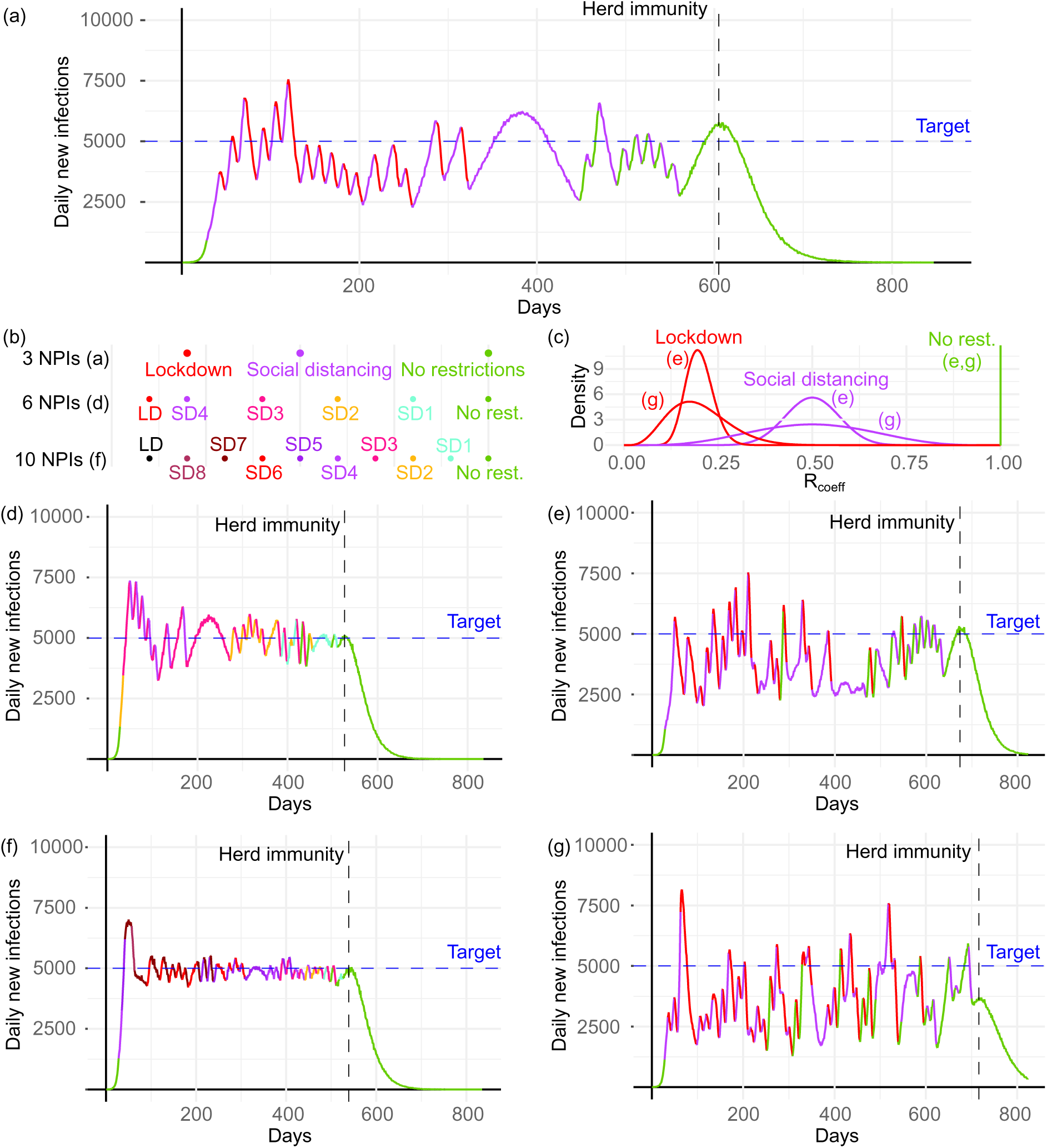
Phased COVID-19 interventions with infection acquired immunity and uncertainty in control efficacies. We explore scenarios for balancing (and easing) a complex NPI space as immunity from infection increases (susceptible individuals are depleted). Scenarios are coloured by interventions chosen. We assume a constant population of 3.5 million individuals and COVID-19 disease characteristics (from **Figs 2-3**). Panel (a) is our baseline with 3 options (no restrictions, social distancing, lockdown) that have fixed (known) effect. We use EpiControl to stabilise infections around the target and minimise costs. Herd immunity is attained at the vertical line (this is not known to EpiControl). Panel (b) illustrates alternative, complex action spaces i.e., a range of intermediate interventions, while panel (c) considers uncertainty over intervention effects as distributions over the coefficients that determine how much they change reproduction numbers. Here, SD refers to social distancing (numbered by stringency, e.g. SD3 is a stronger control than SD2). LD refers to lockdown and represents our most stringent intervention. Panels (d) and (f) demonstrate how having 6 and 10 control choices, respectively, improves on (a). EpiControl solves these more difficult optimisation problems and finds finer-grained policies that leverage the larger intervention space and the rising infection-acquired immunity to stabilise the epidemic more tightly around our target, while gradually reducing intervention stringency and cost. The exit strategy it proposes is phased, optimal and lifts all controls at a timepoint coincident with reaching herd immunity. Panels (e) and (g) show how stochasticity and uncertainty in the effects of the NPIs in (a) alter performance. Every time a measure is enacted, its effect is sampled from a distribution in panel (c). Uncertainty about control is moderate in (e), whereas in (g) it is large, indicating limited knowledge about the intervention impacts. Optimal performance necessarily deteriorates as uncertainty elevates (the controlled epidemic trajectories are more variable) but EpiControl still stabilises infections around the desired target, providing policy recommendations that are robust.

These improved policies offer optimal phased exit strategies that align with the Roadmap [44] of case study 2. Importantly, these scenarios are flexible and easily tested against alternative policy priorities and NPI space counterfactuals, a point that was integral to the Roadmap in which differing numbers of preset steps for lifting lockdown were assessed [40]. Interestingly, all policies confirm our expectation of what is an optimal exit: using progressively milder (and less costly) NPIs as the susceptible population shrinks. These results suggest that larger NPI spaces will yield better results. However, there is a key trade-off in increasing the policy space since uncertainty in NPI effects (i.e., the *c* coefficients) is a strong limiting factor, meaning that the value of having many options may not always be realised. We demonstrate the influence of this uncertainty in **Fig 5e** and **Fig 5f** by including distributions over the effects from our baseline as in **Fig 5c**. Every time a measure is enacted, its effect is sampled from the plotted distribution. We also considered distributions over the costs of NPIs (and these are also easily included in EpiControl) but do not include these results here as their influence was outweighed by the impact of the distributions in **Fig 5c**.

This follows because, under standard unimodal cost distributions where the mean provides an appropriate expected cost, evaluating our candidate interventions across an ensemble of projections largely recovers this expected cost. This differs from uncertainty in intervention efficacies, which shapes transmission dynamics directly as well as nonlinearly and accordingly produces substantially different epidemic trajectories even for the same nominal intervention. Consequently, we focus on uncertainty in NPI efficacy, as this represents the salient source of uncertainty for controlled epidemic dynamics in this setting.

Although fixed effects are convenient for policy planning and frequently (albeit with sensitivity tests) assumed in scenario modelling, they are strong assumptions as behavioural, ecological and other contexts can alter the effect of the same action. EpiControl allows modelling of these variations. We sample lockdown and social distancing effects from the Beta distributions in **Fig 5c** at each implementation, with results for moderate and large uncertainties (broader distributions) in **Fig 5e** and **Fig 5f** respectively. Hence the same nominal intervention may have different realised effects across time. We do this when simulating the ground truth and when projecting to calculate the value function at policy revisions. The no restrictions option always has *c=1*. We find that as uncertainty increases, fluctuations around the target amplify but EpiControl is still able to robustly control the epidemic and finds exit strategies that retain the property of gradual relaxation. This robustness complements that found in case study 2, confirming the benefits of our feedback-learning framework. The flexibility of EpiControl for testing probabilistic effects on complex intervention spaces provides a principled basis for exploring and supporting policy decision trade-offs under real-world uncertainty.

Exploring and optimising over a larger action or decision space with uncertainty is necessarily a more computationally complex problem. We therefore also benchmark the trade-off between the complexity of the simulated scenarios and the computational effort required by EpiControl in the Supplementary Material (see **Fig S3** and **S4**). There we investigate how computational run time changes with key dimensions of problem complexity, including the size of the action space, the policy review frequency, the prediction window length and the simulation ensemble size. Across a wide range of settings and problem parameters, we report the median running time and find approximately linear complexity with maximum runtimes under 1 hour. Hence EpiControl remains well suited for identifying optimal policies in real time and scales well with the sophistication of problems that are likely to interest public health modellers.

## Discussion

In this paper, we have introduced EpiControl, a novel tool that bridges the critical gap between real-time response and using principled algorithms to optimise and update those responses for supporting policy decisions. Across empirical and simulated case studies we demonstrated how EpiControl balances conflicting objectives such as limiting hospital burden and societal closures, to find solutions that evade more traditional approaches. This was exemplified in our analysis of the threshold control scheme proposed in the influential COVID-19 Report 9 [11] (case study 1). EpiControl automatically discovered rules for implementing and relaxing NPIs that reduced both total and peak ICU cases without increasing the duration (and hence cost) of those NPIs. This not only outperformed the threshold scheme but also revealed alternative optimal policies that could trade-off reducing peak ICU cases with fewer NPI actions. This is a key benefit of EpiControl: it helps map the space of optimal decisions based on explicit policy priorities, supporting more transparent and tailored real-time decision-making.

Because EpiControl combines error-correcting feedback against policy targets and adaptive learning of transmissibility parameters, its solutions remain robust and automatically update as new data become available in response to unanticipated shifts such as emerging variants, waning immunity or sudden changes to transmissibility. This was epitomised by our study of the impactful Roadmap out of Lockdown [44] (case study 2). Using empirical data, EpiControl produced optimal scenarios that were consistent with the Roadmap when model assumptions held and as vaccinations were rolled out (though EpiControl used no vaccine data). However, unexpected emergence of the highly contagious Delta variant invalidated those assumptions and meant that traditional scenario models had to be refit under revised conditions [39]. In contrast, EpiControl dynamically adjusted its policies as estimated transmissibility varied, implicitly accounting for the impact of Delta without explicit model revision and proposing optimal exit strategies out of lockdown that still indirectly factored in vaccination and remained effective and reliable. This exemplifies how EpiControl can enable adaptation to unmodelled disease drivers and improve how we generate and update counterfactuals in the unpredictable landscape of disease.

EpiControl addresses two important gaps in how models inform policymaking. First, recent studies recognising the limitations of complex models and a lack of rigorous scenario planning tools have recommended the integration of formal decision analysis frameworks [4], [5], [59].

The feedback and reinforcement learning methodology that underpin EpiControl are examples of decision frameworks that have been successful in machine learning and control engineering applications but have rarely been used in infectious disease epidemiology. This convergence opens a pathway for better integrating artificial intelligence advances into public health policy design [60]. Second, reviews comparing models of differing complexity show that although their forecasts may diverge, they often agree on the relative effectiveness of interventions [15], [16], [61], [62] Hence, parsimonious models can rapidly generate optimal strategies that align with more detailed analyses in terms of decision-relevant outputs, underscoring the value of approaches like EpiControl that maximise the informativeness of simple models.

However, EpiControl cannot replace (nor is it meant to) detailed epidemiological modelling. In favouring simplicity and real-time performance, it necessarily has limitations. Specifically, it assumes homogeneous population mixing and transmission characteristics, ignoring contact heterogeneity and differences in susceptibility or infectivity, all of which may be important for informing targeted interventions. While including this level of detail tends to preclude real-time response, we aim in future to succinctly embed some heterogeneities using metapopulation models [63]. Additionally, our tool assumes target objectives are clear and that efficacies and costs can be specified (at least relatively). Although these may not be known accurately, this requirement can be a strength as EpiControl compels users to make objectives explicit and to integrate the best available information on the costs and benefits of interventions, encouraging transparent, goal-driven decision-making even under uncertainty [4], [62].

We also note that although the social-distancing interventions considered here are illustrative, they translate to NPIs of different intensity or scope in real-world settings [64]. Furthermore, whilst we focussed on case studies where the simulation model is relatively lightweight to support real-time decision-making, EpiControl is designed to be modular and extensible. For example, it can help optimise not only the timing and strength of interventions, but also their spatial scale, provided that the transmission model and action space are defined over relevant spatial units. This would allow spatially targeted control policies to be commonly evaluated under a unifying MPC framework, with interventions selected according to their projected ability to meet epidemiological targets while minimising intervention costs.

Given this context, EpiControl offers distinct advantages for real-time policy support. It uses routinely available disease surveillance data, has roughly linear computational scaling and enables users to customise and stress-test control scenarios rapidly. Case studies 3 and 4, which established how EpiControl can quantify the value of different information streams for guiding actions and optimise complex decision spaces under realistic uncertainty, illustrate its broad functionality. While we focused primarily on uncertainty in intervention efficacy and from leveraging different data streams, uncertainty in the observation processes, specifically under-reporting and reporting delays, may also affect the timing and reliability of control decisions. Some of these effects have been investigated in previous work [25] and provide useful context for more widely interpreting optimal performance amidst the various interacting uncertainties in real-world epidemic settings that must be negotiated for real-time response.

Importantly, by emphasising explicit policy targets that can be updated in real time, EpiControl keeps expert human judgment at the centre of its algorithms. Its plug-and-play design with estimator, predictor, and evaluator modules, allows seamless integration with estimates from tools like EpiEstim (see online vignettes), forecasts from dedicated hubs [26], [61] and cost-benefit analyses from detailed economic-epidemiological models [11], [32]. Consequently, EpiControl can complement and validate more detailed decision models, or emulate them to efficiently explore parameter spaces and improve calibration under uncertainty [59], [65], [66]. We envision EpiControl as the first step to increasing automation in the generation of model-based evidence for epidemic decision-making, synergising advances from engineering and artificial intelligence with well-established epidemic modelling to support modern human-in-the-loop outbreak preparedness pipelines and help decision-makers act confidently and quickly, even when faced with the uncertainty of a disease X [67].

## Methods

We outline the core methodology underpinning the EpiControl results of the main text. These key functionalities together with several flexible options are implemented as a user-friendly R package. This is openly available at https://github.com/sandorberegi/EpiControl with vignettes reproducing all 4 case studies at https://sandorberegi.github.io/EpiControl/. There we provide additional vignettes on how to input Ebola virus disease data, tune algorithm hyperparameters using Bayesian optimisation and integrate EpiControl with outputs from popular estimation tools like EpiEstim [19]. The modular structure from **Fig 1** allows easy integration with existing workflows or swapping of modules for preferred models.

### Renewal models with changing interventions

We model the spread of the pathogen in the population as a renewal branching process. This is a widely used framework in infectious disease epidemiology for estimating transmissibility and forecasting upcoming infections [19], [68]. The renewal model links the incidence of new infections, *I_t_* at present time *t*, to past infection incidence (times *x* < *t*) as in **Eq. (1)**, with Pois indicating Poisson noise and parameter *R_t_* as the time-varying, effective reproduction number.

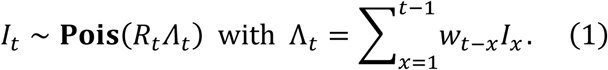

Here Λ*_t_* is the total infectiousness of the disease and is a weighted sum of the past incidence. The weight *w_t_*_−*x*_ is the probability that an infection is transmitted in *t* − *x* time units and the set of weights compose the generation time distribution of the disease with 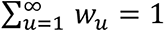 [69].

The generation time distribution completely defines the transmission intervals of the disease and is usually estimated from additional (independent) transmission interval data.

A critical part of modelling interventions is realistically incorporating how they impact the time-varying reproduction number *R_t_* over time. We make the standard assumption that a control action or intervention reduces transmission by a multiplier or coefficient *c* [22], [23]. Hence *R_t_* = *cR*_0,*t*_, with *R*_0,*t*_ as the baseline reproduction number. We allow *R*_0,*t*_ to vary in response to exogenous changes in transmission (e.g. emerging variants or changing immunity). When there are no exogeneous factors and most of the population is susceptible to infection then *R*_0,*t*_ equals the basic reproduction number *R*_0_. The multiplier *c* describing any intervention can be fixed or probabilistic to model realistic variations and uncertainties. For the latter option, we sample realisations of *c* from a Beta prior distribution at every policy review time.

When an intervention is applied or changed, EpiControl offers 3 options for simulating how the old intervention denoted as *R_t_* = *R*_old_ (when there is no intervention *R*_old_ = *R*_0,*t*_) transitions to the new proposed action state *R_t_* = *R*_new_. The first option applies a step change at time *t*_change_ as in **Eq. (2)**. This matches [22] and models instantaneous interventions.

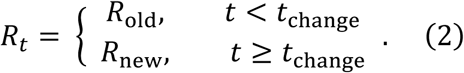

The second option considers gradual changes in transmissibility by embedding the influence of control over the infectious period of individuals that are actively transmitting. This alters the renewal model as in **Eq. (3)** and models interventions as impacting cohorts of infections [70].

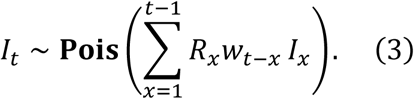

This provides a smoother change from *R*_old_ to *R*_new_ i.e., if the new intervention is enacted at time *t*_change_, then *R_x_*_<*t*change_ = *R*_old_ and *R_x_*_≥*t*change_ = *R*_new_. This option reflects that control often takes time to become established and is unlikely to affect infections equally and at once.

The last option is an alternative approach to modelling how intervention effects disperse or accumulate over time. We apply a logistic transition as in **Eq. (4)**, with θ as a customisable parameter modulating the steepness of the transition.

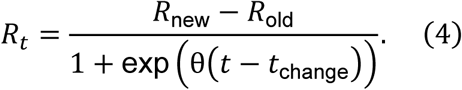

Different choices of θ induce slower or faster transitions between interventions after a policy change. Larger θ leads to sharper transitions, with *θ* → ∞ recovering the step change of option 1. These options cover the range of possibilities when modelling interventions and offer flexible ways of managing the dynamics of changing transmission.

As interventions occur against a background of changing susceptibility, we allow decomposing the impact of interventions from those of susceptibility as *R_t_* = *cR*_0,*t*_*S_t_N*^−1^. Here 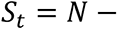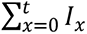 is the number of susceptibles in the population (assuming an infected cannot become susceptible again over the timescale modelled), *N* is the total population size and *R*_0,*t*_ is the baseline reproduction number. If information about susceptibility is available, we can include the influence of infection-acquired immunity, vaccination programmes and waning immunity by adding other multipliers to *S_t_*. In the absence of this information, we can simply apply the formula above, in which case *R*_0,*t*_ bundles unknown effects. However, the updating and rolling optimisation of our algorithm means it performs robustly despite these unknowns. In some simulations we neglect susceptible depletion by setting *S_t_* ≡ *N* and *R_t_* = *cR*_0_.

### Surveillance noise and reporting imperfections

Since infections are rarely observed, to simulate realistic surveillance settings we provide an option in EpiControl to add reporting noise, modelling the observation of case counts or other proxies for new infections [71], [72]. We generalise the renewal models above to output the incidence of symptomatic cases, *C_t_* at time *t*. We consider two central sources of surveillance noise, reporting delay and under-reporting [46]. Reporting delay is the lag from infection onset to case reporting. For a single case, we assume this delay follows a Gamma distribution τ ∼ **Gam**(α_lag_, β_lag_), with shape and scale factors α_lag_ and β_lag_ respectively. Cases *C*̃*_t_* reported on day *t*, subject to reporting delays only, follow the additional Poisson distribution in **Eq. (5)**.

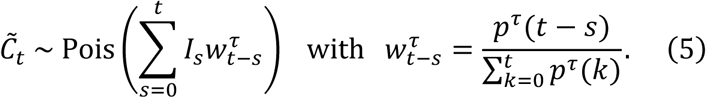

The weights 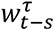 are derived from the Gamma distribution above, which has distribution *p^τ^*. We also include case under-reporting, which arises because not all infections are reported as cases. For example, asymptomatic and mildly symptomatic infected individuals are unlikely to ever get tested or seek treatment. We model this via the Beta-binomial distribution in **Eq. (6)**.

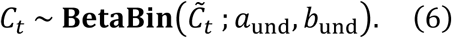

Here *a*_und_ and *b*_und_ respectively parametrise the Beta reporting distribution. In some cases, we may choose to simulate epidemics with only one surveillance noise type or without noise. EpiControl provides the option to turn reporting delays off, leading to *C*̃*_t_* = *I_t_*, or to turn under- reporting off, resulting in *C_t_* = *C*̃*_t_*. For perfect surveillance (no noise), *C_t_* = *C*̃*_t_* = *I_t_*.

### Optimal intervention scenarios using adaptive control and learning

The core of the EpiControl package is an adaptive optimisation algorithm that adapts theory from model predictive control and reinforcement learning [35]. These are overlapping methods for designing optimal policies amid uncertainties. Our algorithm balances 2 major conflicting goals of policymakers: i) minimising undesirable health outcomes (e.g., reducing incidence to a level that prevents hospital capacities or resources from being overloaded) and ii) minimising economic and other costs (e.g., societal disruption) of the interventions chosen to achieve i). We encode these goals in a cost function *ψ*(*t*) as in **Eq. (7)**. This function can be customised to different policy priorities that reflect the local context of the epidemic or region under study.

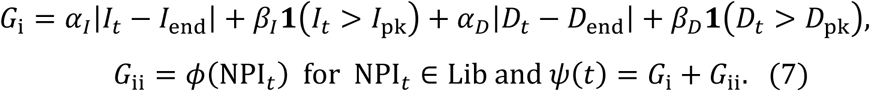

Here *G*_i_ reflects goal i) by penalising deviations in infection incidence *I_t_* and the incidence of deaths from the disease *D_t_* from target endemic levels (*I*_end_, *D*_end_) that policymakers deem to be manageable. Additional penalties are included to prevent large overshoots in these signals from a target peak (beyond which maybe capacities or resources are overloaded) (*I*_pk_, *D*_pk_).

These targets can be modified by users and additional targets (e.g., on oscillations or epidemic duration) introduced as desired. Choices of the weights (*α_I_*, *α_D_*, *β_I_*, *β_D_*) define the prioritisation among these features. These health outcomes are balanced against *G*_ii_, which embodies goal ii) by assigning costs via a function *φ*(.) for choosing an intervention from among a library of possible actions at time *t*, Lib (e.g., this may include stay-at-home orders, social-distancing, closures and other measures). Usually more stringent control actions carry larger *φ*(.) values. The relative costs and weights can be drawn from the literature (e.g., taking uncertainties and effects from [22], [23], [32]) or simply set by users to explore possible counterfactual scenarios. With this framework EpiControl forces explicit definitions of policy goals, improving model transparency and exposing the limits of epidemic controllability when goals are unrealisable.

Our adaptive algorithm proposes optimal control actions from within our library or action space Lib by assessing the long-term value of those actions. The value *V_t_* of an action, as in **Eq. (8)**, is the discounted cost of its consequences over projections that assume that action is taken.

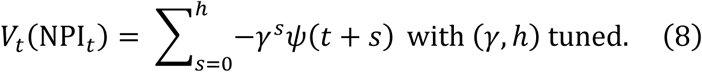

We consider all possible actions from Lib and choose that maximising *V_t_* and hence balancing our conflicting goals i)-ii) over horizon ℎ of interest. This value optimisation process is repeated at every policy review time *T*_rew_ i.e., whenever mod(*t*, *T*_rew_) = 0. This embeds adaptability to changes or uncertainties and offers the flexibility to update costs, effects and priorities in real-time. As it is neither feasible nor desirable to change policy frequently, we set *T*_rew_ to be much longer than our time-step (e.g., here we model daily spread with *T*_rew_ of the order of weeks), but this is customisable. At every *T*_rew_, we re-assess and re-optimise the applied policy.

Both horizon ℎ ≥ 1 and the discount factor *γ* ≤ 1 determine the importance of longer-term versus immediate costs (smaller *γ* or ℎ prioritises short-term costs) and are hyperparameters that EpiControl tunes with Bayesian optimisation algorithms [73], [74]. Tuning is essential, as in all reinforcement learning approaches, because hyperparameters shape the value function and hence optimal actions. Our tuning approach searches for hyperparameter choices that maximise realised long-term performance by applying candidate actions and evaluating their impact from deployment until the simulation end *t*_end_. For diseases with generation times of a few weeks, we set *t*_end_ to several hundred days. We solve our search problem by minimising the undiscounted value function *V*_tune_ of **Eq. (9)** over an ensemble of *m* simulations.

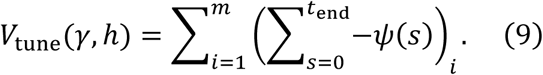

The resulting hyperparameter values ensure that EpiControl, during real-time optimisation of **Eq. (8)**, is appropriately balancing short-term adaptivity with long-term control effectiveness. Because *V*_tune_ is stochastic and computationally expensive, requiring *m* complete epidemic simulations for each query, we use Bayesian optimisation with a Gaussian process surrogate model. The Gaussian process acts as a kernel-based emulator that provides a mean estimate and an uncertainty measure from limited observations, allowing efficient exploration of the constrained parameter space. This enables EpiControl to identify hyperparameter settings that are near optimal with relatively few costly evaluations and allows our framework to be easily adapted by users for an array of diseases and intervention optimisation problems.

When projecting the epidemic dynamics to solve **Eq. (8)**, we assume that the ‘ground truth’ and our model for projections follow the same renewal branching process model from **Eq. (1)** with the same generation time distribution. However, we re-estimate the reproduction number from unfolding data, to respond to unfolding changes in transmissibility (e.g. from susceptible depletion). This can be thought of as the adaptive control version of the real-time estimation procedures that are widely used to track epidemics (e.g., EpiEstim). We estimate reproduction numbers using the left expression in **Eq. (10)**, which is based on the formula proposed in [19].

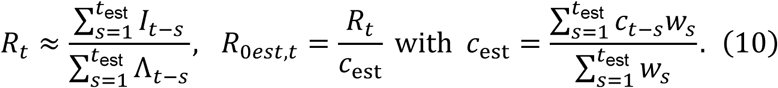

Here *t*_est_ is a chosen estimation time window reflecting the autocorrelation of transmissibility parameters. EpiControl allows for replacing this estimation procedure with other popular *R_t_* estimation packages including EpiEstim (our package directly interfaces with this approach, and we provide a vignette on its use) and EpiFilter [75]. We also need to estimate the baseline reproduction number *R*_0*est*,*t*_ by considering the context of any existing NPIs at that time and their impact on transmissibility. Consequently, we further infer an intervention efficacy *c*_est_ as the right expression in **Eq. (10)**, which considers the historical impact of our policies over *t*_est_.

When surveillance is noisy, we replace *I_t_* with the incidence of cases *C_t_* and apply appropriate delay and under-reporting parameters from the literature. We may also use the incidence of deaths *D_t_* or ICU cases instead as these can also be modelled as under-ascertained, delayed proxies of *I_t_* [46]. Policy goals can similarly be updated to reflect these signals. Users can also tailor their choice of observed signal based on which is most reliable for the region under study (e.g., if case reporting is highly behaviour-dependent, then ICU cases may be more reliable despite their larger delays). EpiControl also allows replacing the models for these signals. As an example, we can swap the above renewal models for agent-based simulators to generate a ‘ground-truth’ that captures population heterogeneities or use different generation times for the ground truth and projection modules to incorporate the effect of model mismatch.

### Exogenous changes in transmission: co-circulating variants and vaccination

EpiControl can simulate exogenous or environmental changes such as emerging variants or vaccination protocols. We note that whilst we offer the functionality to also incorporate these dynamics within optimisations, our projections by default do not directly use this information. Nevertheless, the algorithm adapts through periodic re-estimation of the reproduction number and re-evaluation of the value function.

We model how the baseline and basic reproduction numbers shift as an emerging pathogenic variant dominates the currently circulating variant in **Eq. (11)**.

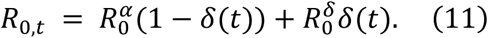

Here *δ*(*t*) is the prevalence of the emerging variant i.e., the fraction of total active infections that it contributes and is a signal that is exogeneous to our simulations. Superscripts *α* and *δ* denote the original and emerging variants. We do not directly model any interactions between variants (though this is a planned extension of our package), but we include the impact of such interactions implicitly within the linear mixing model of **Eq. (11)**. If we do not have prevalence information then EpiControl acts under the models described above and the effect of mixing is an unknown, implicit to *R*_0,*t*_. This is also how we handled unknown population susceptibility earlier i.e., unknowns affecting transmissibility are bundled into the estimated baseline *R*_0*est*,*t*_.

Last, we include the option of modelling vaccination campaign effects (even with co-circulating variants) within EpiControl. We let *v*(*t*) be the proportion of the population that is vaccinated (another exogenous variable) with *σ^α^* and *σ^δ^* as the vaccine efficacies against the circulating and emerging variants. **Eq. (12)** then describes an overall vaccine protection factor *η*(*t*).

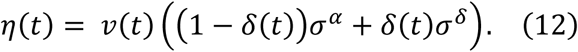

## Supporting information

Supplementary analyses

## Data and code availability

All data and code to reproduce this study are open-access and available in the EpiControl package https://github.com/sandorberegi/EpiControl with vignettes including additional use-cases covering Ebola virus disease data, Bayesian optimisation for hyperparameter tuning and interfacing with tools such as EpiEstim [19] at https://sandorberegi.github.io/EpiControl/.

## Author Contributions

Conceptualisation: K Parag. Investigation: S Beregi and K Parag. Methodology: S Beregi and K Parag. Formal analysis: S Beregi. Funding acquisition: K Parag. Software: S Beregi and S Bhatia. Supervision: K Parag. Validation: S Beregi, A Cori and K Parag. Visualisation: S Beregi, A Cori and K Parag. Writing – original draft: S Beregi and K Parag. Writing – review & editing: all authors.

This protection factor captures the mixing of efficacies due to the mixing of variants and changes the baseline reproduction by adding a multiplier (1 − *η*(*t*)). If this is known, similar to the variant and susceptibility cases we can extract this effect. If unknown (as in case study 2), this factor is again implicitly encoded in our baseline *R*_0*est*,*t*_. The EpiControl algorithm adapts its optimal control solutions to these unknowns by re-estimating *R*_0*est*,*t*_ at each policy review.

## Data Availability

All data and code to reproduce this study are open-access and available in the EpiControl package https://github.com/sandorberegi/EpiControl with vignettes reproducing all our case studies and introducing additional functionality at https://sandorberegi.github.io/EpiControl/.

