## Supplementary analyses for "EpiControl: a data-driven tool for optimising epidemic interventions and automating scenario planning to support real-time response"

We introduce additional simulations (figures S1-S4) to benchmark and test the sensitivity of the EpiControl package and its application within our case studies. Further analyses and examples are also available online at https://sandorberegi.github.io/EpiControl/.

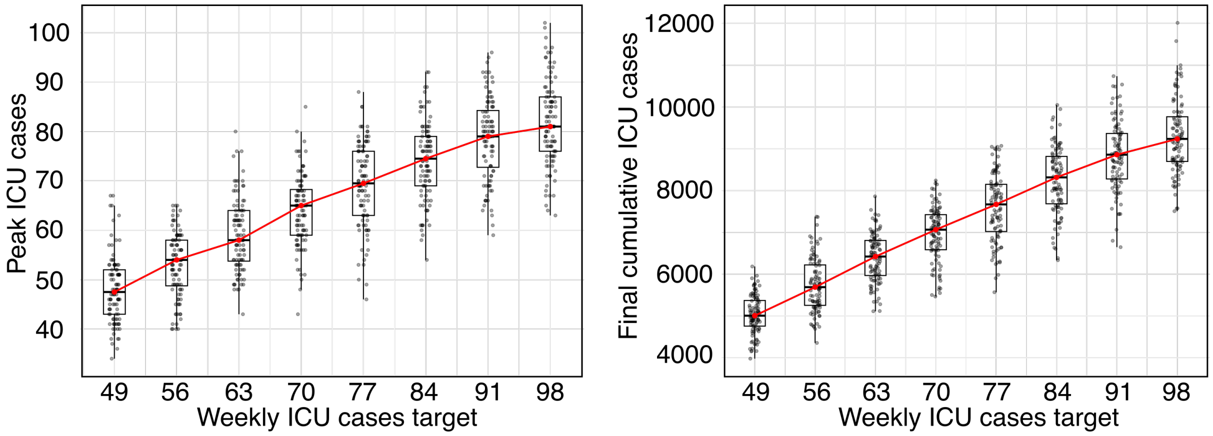


**Figure S1. Target-sensitivity when controlling COVID-19 ICU cases.** We benchmark the performance robustness of Case Study 1 (see **Fig. 2** of the main text), by assessing sensitivity to the chosen ICU target. Boxplots show outcome distributions from 100 stochastic simulations for each ICU target value. The left panel shows peak ICU cases, while the right panel shows final cumulative ICU cases. In each boxplot, the box shows the middle 50% of simulated outcomes, the horizontal line shows the median, and the whiskers extend to the most extreme values within 1.5 times this middle-50% range. The red line connects the median values across target settings. Median outcomes increase approximately linearly with the target, indicating that small changes in the ICU target do not produce markedly different outcome distributions. Relative variation is larger for the peak ICU cases because cumulative outcomes combine cases over time and therefore smooth random fluctuations.

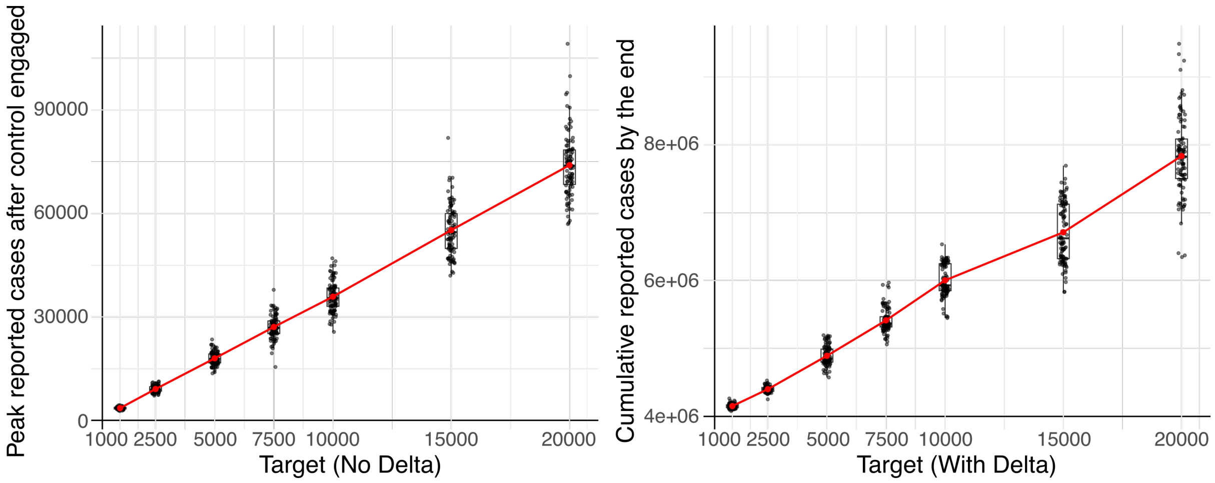


**Figure S2. Target sensitivity in the COVID-19 vaccination case study.** We test the sensitivity of epidemic outcomes to the incidence target for COVID-19 vaccination scenarios from Case Study 3 that exclude and include the unknown emergence of the Delta variant (see **Fig. 3** of the main text). Left panel: final cumulative reported cases in the scenario without Delta. In this case, vaccination eventually achieves herd immunity and epidemic elimination, making final epidemic size an appropriate outcome measure. Right panel: peak reported cases for the Delta-variant scenario, measured after the simulation has started and incidence has first been brought close to the target. In this scenario, immunity is driven by vaccination, but vaccination alone is insufficient to bring the effective reproduction number below 1 without interventions. As shown in **Fig. 3**, herd immunity and elimination are not achieved, but instead there are sustained oscillations of infections around the target. Boxplots illustrate the distribution across stochastic simulations for each target. The box covers the 25-75% range with the central horizontal line giving the median and whiskers extending to extreme values. Individual points show simulation outcomes, and the red line connects the mean outcome at each target value. In both panels, the mean outcome increases approximately linearly with the target. Peak-case distributions in the Delta scenario are broadly unimodal with similar dispersion across targets. In the no-Delta scenario, final cumulative cases show some bimodality at intermediate targets, such as around 5,000, 7,500 and 10,000. This is consistent with the relatively small number of policy switches in the optimal strategies, where small stochastic differences can lead to distinct but similar control trajectories, as also seen in **Fig. 3**. The horizontal axes in all panels are linear but labelled to highlight some key values.

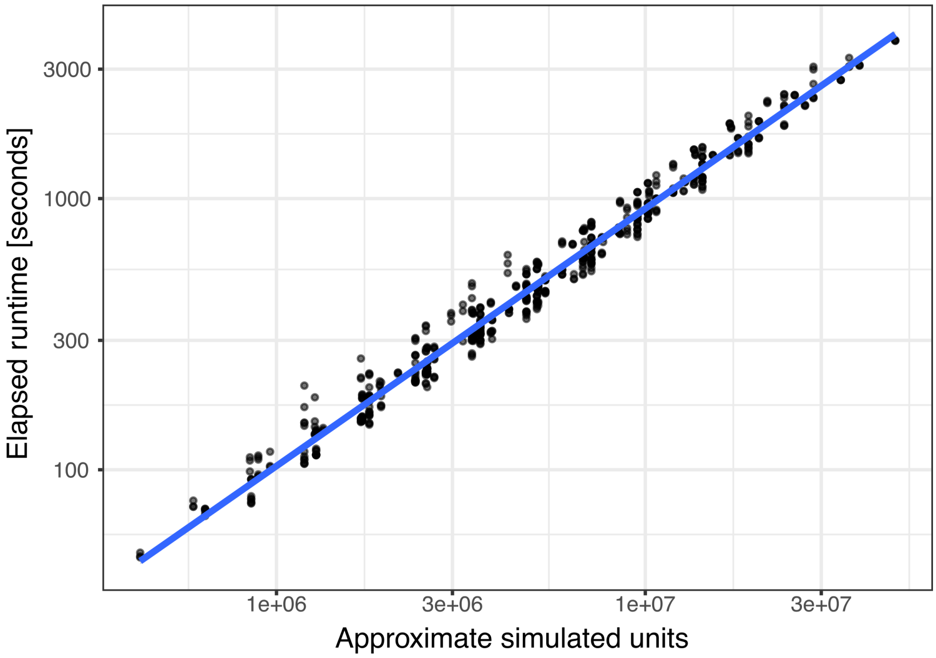


**Figure S3. Runtime scaling with total MPC simulation burden.** We examine computational complexity for the EpiControl MPC algorithm. These benchmarks use the stochastic the renewal-model epidemic-control setup detailed in **Fig. 1** and the Methods. We use COVID-19 epidemiological parameters ($R_0$ = 3.5, a generation time distribution with mean 6.5 days and variance of 2.1) and surveillance noise values (reporting delays with mean 10.5 days and variance 5.0 and under-reporting with mean reporting probability 0.3) and neglect susceptible depletion. In this setup, the controller repeatedly selects among interventions that reduce transmission, with the objective of keeping incidence close to a target of 5,000 cases while penalising overshoots and costly control actions. Every point represents a distinct benchmarking configuration of this procedure, run locally on a MacBook Pro (2.9 GHz 6-Core Intel Core i9, 32 GB RAM, OS: Sequoia 15.7.5). The horizontal axis summarises the total computational burden implied by these configurations. We calculate this proxy for problem complexity as Action space size × Prediction horizon × Simulation ensemble size × Number of policy reviews, where the number of policy reviews is given by the total simulation length divided by the policy review period. The vertical axis shows the corresponding runtime. The near-linear relationship on the log-log scale (the gradient is approximately 1) indicates that computation time is driven by the number of forward simulations or evaluations required, with no evidence of abrupt changes or computational blow-up across the tested range. The MPC implementation scales predictably as the policy optimisation problem becomes progressively more complex. Importantly, every benchmark configuration completed in under one hour, supporting the suitability of the implementation for real-time or near-real-time policy analysis.

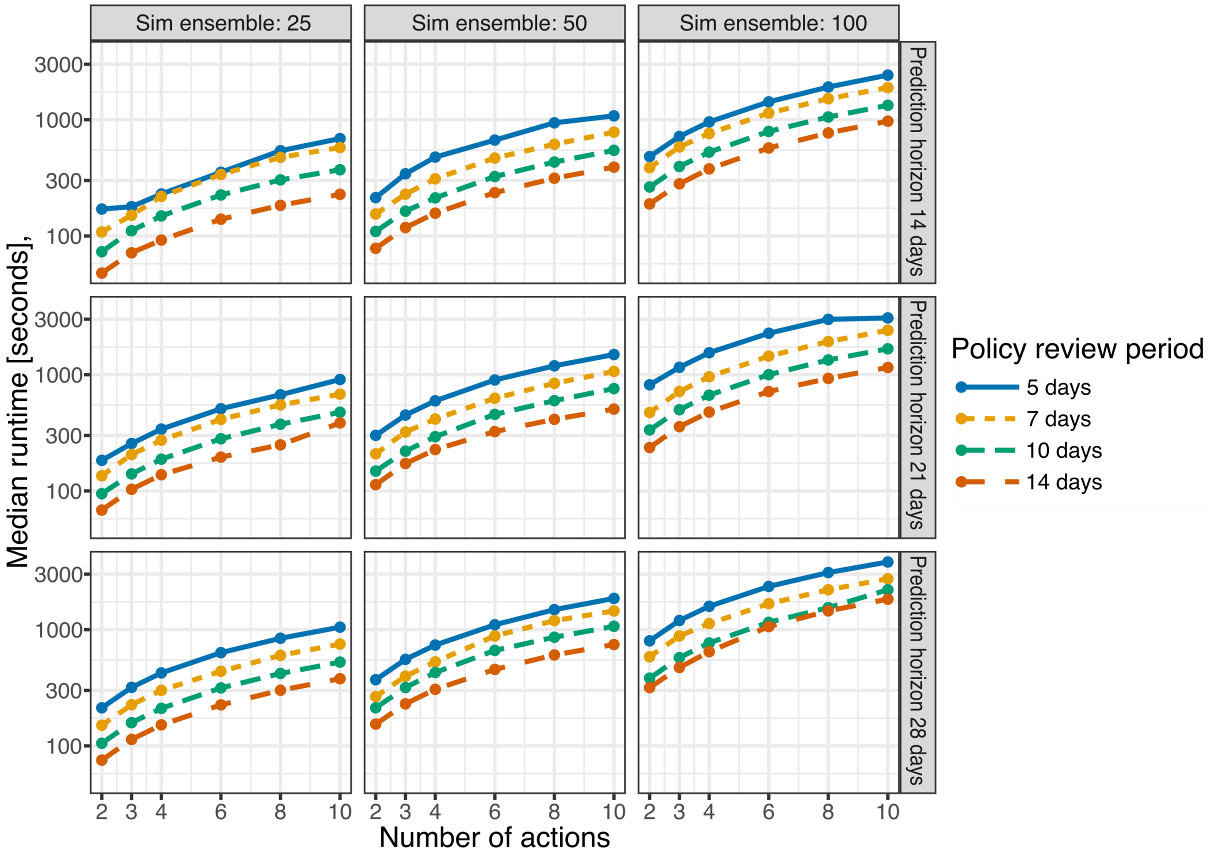


**Figure S4. Runtime sensitivity to individual MPC settings.** We investigate how EpiControl MPC runtime changes with individual optimisation settings, using the same stochastic renewal-model epidemic-control benchmark setup from **Fig. S3**. This setup assumes COVID-19 epidemiological parameters, delayed and under-reported incidence observations, no susceptible depletion, and an incidence target of 5,000 cases. Benchmarking results show how runtime on a MacBook Pro (2.9 GHz 6-Core Intel Core i9, 32 GB RAM, OS: Sequoia 15.7.5) varies with the size of the available action space under different simulation ensemble sizes, prediction horizons, and policy review periods. Columns indicate increasing simulation ensemble sizes, rows show increasing prediction horizons and coloured lines illustrate various policy review periods. Runtime increases with larger action spaces (more decision variables), larger simulation ensembles, longer prediction horizons and more frequent policy review. The 5-day review period is consistently the most computationally demanding, while the 14-day review period is fastest, reflecting its smaller number of policy updates. Across panels, the curves remain smooth and approximately parallel, showing that runtime increases in a regular and interpretable way rather than becoming unstable for larger benchmark settings.
